# Spatial prediction of air pollution levels using a hierarchical Bayesian spatiotemporal model in Catalonia, Spain

**DOI:** 10.1101/2021.06.06.21258419

**Authors:** Marc Saez, Maria A. Barceló

**Affiliations:** Research Group on Statistics, Econometrics and Health (GRECS), University of Girona, Girona, Spain; CIBER of Epidemiology and Public Health (CIBERESP), Madrid, Spain

**Keywords:** Spatial predictions, Hierarchical Bayesian spatiotemporal model, Stochastic Partial Differential Equations (SPDE), Integrated Nested Laplace Approximations (INLA).

## Abstract

Our objective in this work was to present a hierarchical Bayesian spatiotemporal model that allowed us to make spatial predictions of air pollution levels in an effective way and with very few computational costs.

We specified a hierarchical spatiotemporal model, using the Stochastic Partial Differential Equations of the integrated nested Laplace approximations approximation. This approach allowed us to spatially predict, in the territory of Catalonia (Spain), the levels of the four pollutants for which there is the most evidence of an adverse health effect.

Our model allowed us to make fairly accurate spatial predictions of both long-term and short-term exposure to air pollutants, with a low computational cost. The only requirements of the method we propose are the minimum number of stations distributed throughout the territory where the predictions are to be made, and that the spatial and temporal dimensions are either independent or separable.

**Highlights:** We show a hierarchical Bayesian spatiotemporal model.

Our model provides predictions of both long-term and short-term exposure.

The computational cost is low.

The model only needs a minimum number of stations being distributed throughout the territory.

The other requirement of our model is that the spatial and temporal dimensions are either independent or separable.

**Graphical abstract:** 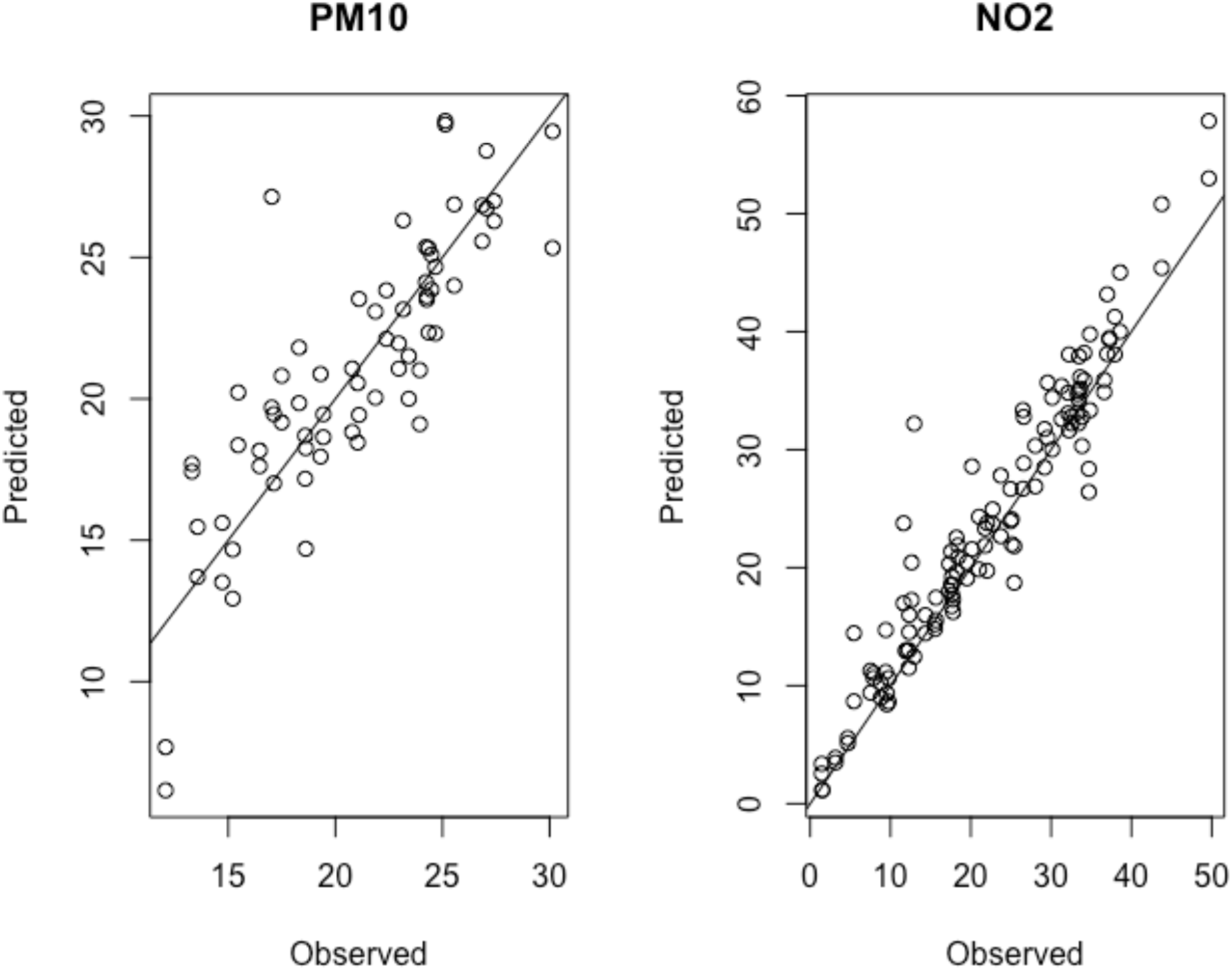

**Software and Data availability:** We used open data with free access using these sources. <u>Air pollutants</u>

Departament de Territori i Sostenibilitat, Generalitat de Catalunya [Available at: https://analisi.transparenciacatalunya.cat/en/Medi-Ambient/Qualitat-de-l-aire-als-punts-de-mesurament-autom-t/tasf-thgu, last accessed on March 14, 2021].

<u>Meteorological variables</u>

METEOCAT, Generalitat de Catalunya. Meteorological data from XEMA [Available at: https://analisi.transparenciacatalunya.cat/en/Medi-Ambient/Dades-meteorol-giques-de-la-XEMA/nzvn-apee, last accessed on March 14, 2021].

AEMET. AEMET Open Data [in Spanish] [Available at: http://www.aemet.es/es/datos_abiertos/AEMET_OpenData, last accessed on March 14, 2021].

<u>Digitized cartography of the ABS</u>

Departament de Salut. Cartography [Available at: https://salutweb.gencat.cat/ca/el_departament/estadistiques_sanitaries/cartografia/, accessed on March 14, 2021].

Code will be available at www.researchprojects.es

## 1. Introduction

In studies assessing the health effects of exposure to air pollution, there is the problem of how to estimate that exposure. Air pollution monitoring station locations do not usually coincide with where the majority of the subjects exposed to such pollution are found. In fact, the air pollution monitoring stations are not often distributed homogeneously in the territory under study, and it is quite usual that large areas, even some densely populated one, do not have any stations at all.

Many studies use the measurements observed in the geographical region of the study to estimate, by means of point estimators, exposure levels for that entire region. The estimators most widely used are the inverse-distance weighted average and the arithmetic mean of the values of the air pollutant observed in several monitor stations, although sometimes the values of the pollutants observed in the nearest monitoring station are also used as estimators. The problem, as Wannemuehler *et al*. (2009) pointed out, is that when air pollution levels exhibit spatial variation across the study region, using these point estimators leads to a bias, as a consequence of ignoring the spatial structure (i.e., spatial dependence) of the data. Furthermore, when that biased estimated level is related to a health variable, this leads to an underestimation of the health effect of interest (Wannemuehler *et al*., 2009).

There are numerous studies that propose models to estimate the levels of air pollutants, explicitly incorporating both spatial and temporal dependence (Cameletti *et al*., 2011, 2013; Pirani *et al*., 2013; Shaddick *et al*., 2013; Liang *et al.,* 2015, 2016; Calculli *et al*., 2015; Cheam *et al*., 2017; Mukhopadhyay and Sahu, 2018; Chen *et al*., 2018; Clifford *et al.,* 2019; Nicolis *et al.,* 2019; Wan *et al.,* 2021) (to refer to only some of those that have appeared in the last ten years). However, we must point out that very few studies attempt to predict air pollution levels in locations where there is no monitoring station (ie, spatial prediction) (Cameletti et al., 2011, 2013; Pirani et al., 2013; Shaddick et al., 2013; Mukhopadhyay and Sahu, 2018; Nicolis et al., 2019), or, having them, to perform out-of-sample temporal predictions (Wan et al., 2021).

The spatial domain of these studies ranges from cities (Santiago de Chile - Nicolis *et al.,* 2019-; Beijing - Wan *et al.,* 2021-) to countries (EU-15 countries - Shaddick *et al*., 2013-), passing through metropolitan areas (Greater London - Pirani *et al*., 2013-) and regions (Po valley, northern Italy - Cameletti *et al*., 2011, 2013-; England and Wales - Mukhopadhyay and Sahu, 2018-). The pollutants that are predicted in these studies are coarse particles, PM_10_, those with a diameter of 10 micrometres (μm) or less (Cameletti *et al*., 2011, 2013; Pirani *et al*., 2013; Mukhopadhyay and Sahu, 2018), fine particles, PM_2.5_, those with a diameter of 2.5 μm or less (Mukhopadhyay and Sahu, 2018; Nicolis *et al.,* 2019; Wan *et al.,* 2021), nitrogen dioxide, NO_2_ (Shaddick *et al*., 2013; Mukhopadhyay and Sahu, 2018) and ozone, O_3_ (Mukhopadhyay and Sahu, 2018). Regarding the frequency at which pollutants are observed, daily data (Cameletti *et al*., 2011, 2013; Pirani *et al*., 2013; Mukhopadhyay and Sahu, 2018) dominate, although hourly data (Nicolis *et al.,* 2019; Wan *et al.,* 2021) and annual data (Shaddick *et al*., 2013) are also used.

The models used in most of these articles, in addition to incorporating spatial and temporal dependencies, include explanatory variables among which appear, in decreasing order of the number of studies, meteorological variables (Cameletti *et al*., 2011, 2013; Pirani *et al*., 2013; Shaddick *et al*., 2013; Nicolis *et al.,* 2019; Wan *et al.,* 2021), other pollutants different from the one predicted (Cameletti *et al*., 2011, 2013), topographical variables (altitude – Cameletti *et al*., 2013; Wan *et al.,* 2021- and distances to sea and roads - Shaddick *et al*., 2013 -, and to mountains - Wan *et al.,* 2021-), site types (Pirani *et al*., 2013; Mukhopadhyay and Sahu, 2018), and land use variables (Shaddick *et al*., 2013).

With one exception (Wan *et al.,* 2021), the studies use a Bayesian approach since it is the one that best allows the uncertainty of complex space-time data to be incorporated. Most of the studies that use the Bayesian approach perform the inference using the Monte Carlo Markov Chain (MCMC) (Cameletti *et al*., 2011; Pirani *et al*., 2013; Shaddick *et al*., 2013; Mukhopadhyay and Sahu, 2018; Nicolis *et al.,* 2019). Only one uses the Stochastic Partial Differential Equations (SPDE) representation of the INLA approximation (Cameletti *et al*., 2013). Using MCMC implies a high computational model complexity that, in some cases, prevents the practical application of the methods proposed by these studies. As an exception, it is worth mentioning Nicolis *et al*. (2019), who use the *spTimer* package (Bakar and Sahu, 2015). This package, which uses MCMC, allows large space-time data sets to be handled with fast computation and very good data processing capacity. The INLA approach is much more computationally effective than MCMC, producing accurate approximations to posterior distributions, even for very complex models (Lindgren and Rue, 2015).

These few studies that provide methods for spatial prediction use a relatively large number of monitoring stations. In this study we intend to present an equally effective model that allows the use of information from a small number of monitoring stations. Furthermore, we intend to make spatial predictions with a computational cost much lower than existing methods.

Specifically, our objective in this work was to present a hierarchical Bayesian spatiotemporal model that allowed us to make spatial predictions of air pollution levels in an effective way and with very few computational costs. In this work, we used the SPDE representation of the INLA approximation to spatially predict, in the territory of Catalonia (Spain), the levels of the four pollutants for which there is the most evidence of an adverse health effect: PM_10_, NO_2_, O_3_ and PM_2.5_. We performed the spatial predictions at a point level (defined by its UTM coordinates), allowing them to be aggregated later in any spatial unit required. We were especially interested in the long-term exposure to air pollutants. That is, by living in a certain area an individual is exposed to a mix of pollutants that have lasting effects on their health. We also considered the performance of our method to spatially predict short-term exposure to air pollutants, which has more temporary effects on health.

## 2. Methods

### 2.1. Data

We obtained information on the hourly levels of air pollution for 2011-2020 from the 143 monitoring stations from the Catalan Network for Pollution Control and Prevention (XVPCA) (open data) (*Departament de Territori i Sostenibilitat, Generalitat de Catalunya*, 2021), located throughout Catalonia (Figure 1), and that were active during that period. The pollutants we were interested in for making spatial predictions were PM_10_, NO_2_, O_3_ and PM_2.5_ (all of them expressed as μm/m^3^) (air pollutants of interest, hereinafter). However, the monitoring stations also measured other pollutants: nitrogen monoxide (NO), sulphur dioxide (SO_2_), carbon monoxide (CO), benzene (C_6_H_6_), hydrogen sulphide (H_2_S), dichloride (Cl_2_), and heavy metals (mercury, arsenic, nickel, cadmium and lead). We have used these other pollutants as covariates.

**Figure 1.**
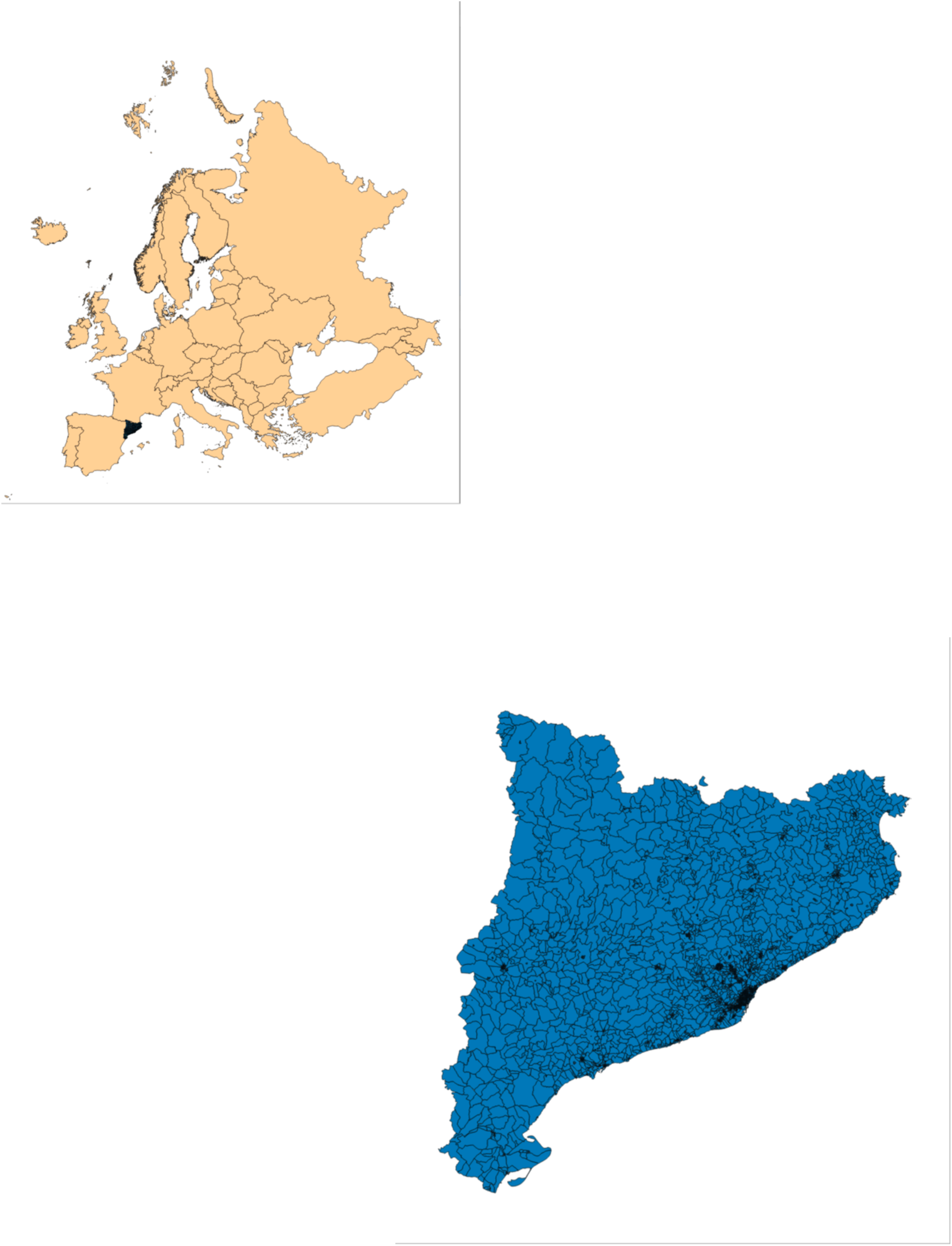
**Location of Catalonia (Spain).**

Not all pollutants of interest were measured at all the monitoring stations. Thus, during the entire period 2011-2020, PM_10_ was measured at 122 stations, NO_2_ at 77 stations, O_3_ at 62 stations and PM_2.5_ at 42 stations. As can be seen in Figure 2, most of the monitoring stations were located in the city of Barcelona and in its metropolitan area. In the rest of the territory, the stations were located in cities (especially those that measure NO_2_ and PM_2.5_) and, in the case of O_3_, also in rural areas. On the other hand, in 2020 (which we used to spatially predict short-term exposure), the number of air pollution monitoring stations dropped considerably, from 143 to 78. In particular, those stations that measured particles dropped dramatically (PM_2.5_ from 42 to 3, 92.88% less; PM_10_ from 122 to 36, 70.49% less). The number of stations that measured O_3_ went from 62 to 50 (19.35% less stations) and NO_2_ from 77 to 67 stations (12.99% less) (Table 1).

**Figure 2.**
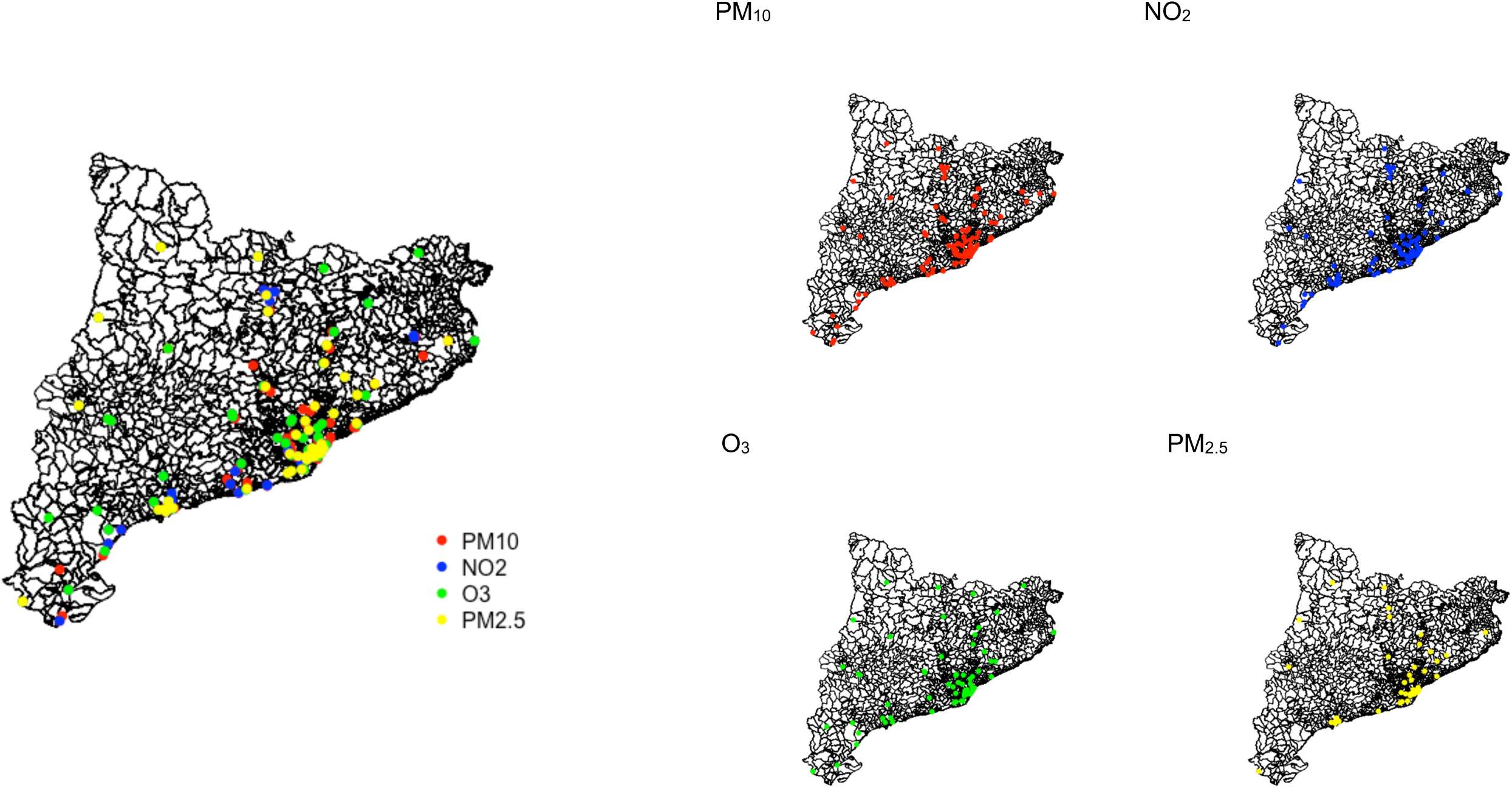
**Distribution in the territory of Catalonia (Spain), of the air pollution monitoring stations, according to where air pollutants (PM_10_, NO_2_, O_3_ and PM_2.5_) are measured.**

**Table 1.** Description of the air pollutants.

|  | Long-term exposure |  |  | Short-term exposure |  |  |  |
| --- | --- | --- | --- | --- | --- | --- | --- |
|  | Number stations | 2011-2018 | 2019 | Number stations | Week 1-36, 2020 | Week 1-10 and 26-36 (excluding lockdown), 2020 | Week 37-48, 2020 |
| **PM<sub>10</sub>** (µm/m<sup>3</sup>)<br>mean (sd)<br>median [Q1-Q3] | 122 | 23.00 (7.79)<br>22.32 [18.10-27.15] | 21.05 (6.37)<br>20.68 [16.67-25.37] | 36 | 19.29 (9.85)<br>17.83 [13.13-23.42] | 21.91 (10.96)<br>20.29 [15.30-26.21] | 20.23 (11.34)<br>18.08 [12.71-25.42] |
| **NO<sub>2</sub>** (µm/m<sup>3</sup>)<br>mean (sd)<br>median [Q1-Q3] | 77 | 25.00 (15.10)<br>23.59 [12.87-35.61] | 21.78 (12.76)<br>20.92 [11.59-30.61] | 67 | 15.49 (12.04)<br>12.25 [6.54-21.46] | 18.40 (13.36)<br>15.21 [7.88-26.46] | 19.07 (12.98)<br>17.11 [8.71-28.13] |
| **O<sub>3</sub>** (µm/m<sup>3</sup>)<br>mean (sd)<br>median [Q1-Q3] | 62 | 53.64 (20.56)<br>55.33 [37.62-68.86] | 55.77 (20.22)<br>57.47 [39.60-70.15] | 50 | 55.23 (19.76)<br>56.67 [43.92-68.00] | 50.90 (21.26)<br>53.27 [36.58-64.79] | 44.55 (19.40)<br>43.92 [30.25-58.42] |
| **PM<sub>2.5</sub>** (µm/m<sup>3</sup>)<br>mean (sd)<br>median [Q1-Q3] | 42 | 13.98 (5.12)<br>13.33 [10.47-16.75] | 10.85 (2.91)<br>10.48 [9.86-11.57] | 3 | 11.42 (7.54)<br>9.33 [5.86-14.51] | 11.94 (8.44)<br>9.33 [5.66-15.34] | 10.30 (6.58)<br>9.13 [5.45-14.02] |
| **Number of stations** | 143 |  |  | 78 |  |  |  |

Daily averages

As we said, our primary interest was in spatially predicting long-term exposure to air pollutants. In this case, we used the monthly averages, after obtaining the daily averages from the hourly data, from January 2011 to December 2019. To make the spatial predictions of the short-term exposure, we used the daily averages from January 1, 2020 to November 29, 2020.

We carried out the spatial predictions at a point level, with the centroids being Basic Health Areas (ABS, for its acronym in Catalan from here on). Catalan health planning defines an ABS as the elementary territorial unit through which primary health care services are organized (*Atenció Primària Girona. Institut Català de la Salut*, 2021). The ABSs are either made up of neighbourhoods or districts in urban areas or by one or more municipalities in rural areas. Their delimitation is determined by geographical, demographic, social and epidemiological factors and, in particular, based on the accessibility the population has to services and the efficiency in the organization of health resources [18]. Catalonia is divided into 376 ABSs, with a population between 371 and 72,321 inhabitants (mean 20,266 inhabitants, standard deviation 13,391, median 18,457 inhabitants, first quartile -Q1- 10,554, third quartile -Q3- 27,529). The population density was in the range of 0.31-34,590.58 inhabitants/km^2^ (mean 3,486.36, standard deviation 6,719.23, median 309.18, Q1 44.83, Q3 3,752.54). In Catalonia, 769 of the 947 of the municipalities belong to a single ABS. Of the 178 remaining, 46 were divided into more than one ABS, 37 of them into a maximum of five ABSs, eight between six and 14 ABSs and one, (the city of Barcelona) into 67 ABSs (Idescat, 2021).

Less than a third of ABSs have at least one air pollution monitoring station (105 from a total of 376). An ABS has five monitoring stations, six ABSs have three stations, 22 ABSs have two stations and the remaining 76 have only one station.

As covariates, we included the altitude of the air pollution monitoring station (in m) and the area of the ABS (in km^2^). The altitude (as well as other information related to the monitoring station, such as its latitude and longitude) were obtained from the *Departament de Territori i Sostenibilitat* (2021). We transformed the geographic coordinates (latitude and longitude) to UTM coordinates (in km) using the R package *rgdal* (Bivand *et al*., 2021). The areas of the ABS, as well as the UTM coordinates of their centroids, were calculated using QGIS (version 2.18) from the digitized cartography of the ABS (information of 2018) (open data) (*Departament de Salut*, 2021).

It is known that, at least in the short term, exposure to air pollution is correlated with various meteorological variables. For this reason, in the case of spatial prediction of short-term exposure, we also included several meteorological variables as covariates. Most of them, such as temperature (in °C), relative humidity (in %), wind speed at 10m (in m/s) and atmospheric pressure (hPA) influence the dispersion of the pollutant; although some also influencing its formation, for instance, global solar radiation (W/m^2^) (O_3_ is a secondary pollutant, formed when the two atoms that make up oxygen gas dissociate under the action of light solar). The sources of the data were the stations of the Network of Automatic Meteorological Stations (XEMA) of the Meteorological Service of Catalonia (METEOCAT) (open data). We also used the daily data from the State Meteorological Agency’s (AEMET) automatic stations.

Albeit not as much as the air pollutants monitoring stations, the meteorological stations are also dispersed throughout the territory. Catalonia has 217 meteorological stations, 188 belonging to METEOCAT and 29 to AEMET. All of them measured all the meteorological variables every day. We used the same model (explained in this work) for short-term exposure to carry out the spatial prediction of the daily values of the meteorological variables at the ABS level, for the year 2020. Further details concerning this, can be found in Ribas *et al*. (2021).

### 2.2. Model specification

We specified a hierarchical spatiotemporal model as follows:

At the top of the hierarchy:

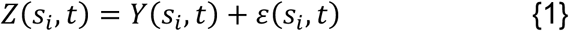

where i denoted the air pollution monitoring station where the pollutant was observed; t was the time unit; *s_i_* was the location of the station; *Y*(. , . ) the spatiotemporal process, the realization of which corresponded to the pollutant measurements (at station i and time unit t); and *ɛ*(. , . ) was the measurement error defined by a Gaussian white-noise process (i.e., spatially and temporally uncorrelated) (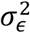 was the nugget effect).

At the next level, we specified the following measurement equation:

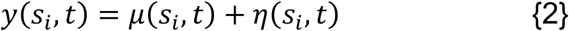

where *y*(. , . ), is the realization of the spatiotemporal process; *μ*(. , . ) denoted the large-scale component, depending on the covariates; and *η*(. , . ) was a spatiotemporal process.

The spatiotemporal process was an independent in time Gaussian field (GF) with zero mean and a Matérn covariance function:

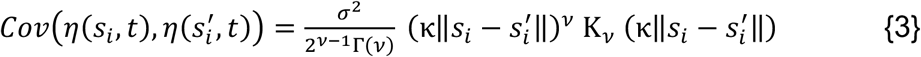

where Κ_ν_ is the modified Bessel function of the second type and order *ν* > 0. *ν* is a parameter controlling the smoothness of the GF, *σ*^2^ is the variance and *κ* > 0, is a scaling parameter related to the range, *ρ*, the distance to which the spatial correlation becomes small. We used 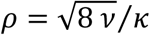, where *ρ* corresponded to the distance where the spatial correlation is close to 0.1 for each *ν* (Lindgren *et al*., 2011). 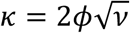, where *ϕ* is a parameter controlling the rate of decay of the spatial correlation as the distance 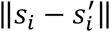 increases.

Due to its computational problems, we chose to represent the GF as a Gaussian Markov Random Field (GMRF) (Rue *et al.,* 2009). GMRFs are defined by a precision matrix with a sparse structure allowing inference to be performed in a computationally effective way. We linked the GF and GMRF through the Stochastic Partial Differential Equations (SPDE) approach (Lindgren *et al*., 2011). The SPDE allowed us to find a GMRF, with local neighbourhood and sparse precision matrix (instead of spatiotemporal covariance function and the dense covariance matrix of a GF, respectively), that best represented the Matérn field. Further details can be found in Lindgren *et al*. (2011) and in Cameletti *et al*. (2013).

We specified the large-scale component, *μ*(. , . ), as a generalized linear mixed model (GLMM) with response from the Gaussian family. Specifically, for each of the pollutants of interest (PM_10_, NO_2_, O_3_ and PM_2.5_) we specified two GLMMs: one for long-term exposure and the other for short-term exposure.

Long-term exposure:

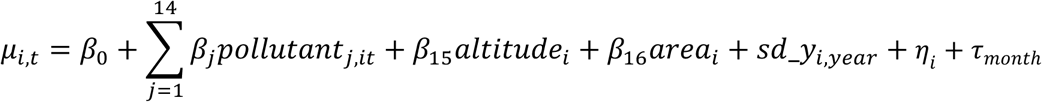

Short-term exposure:

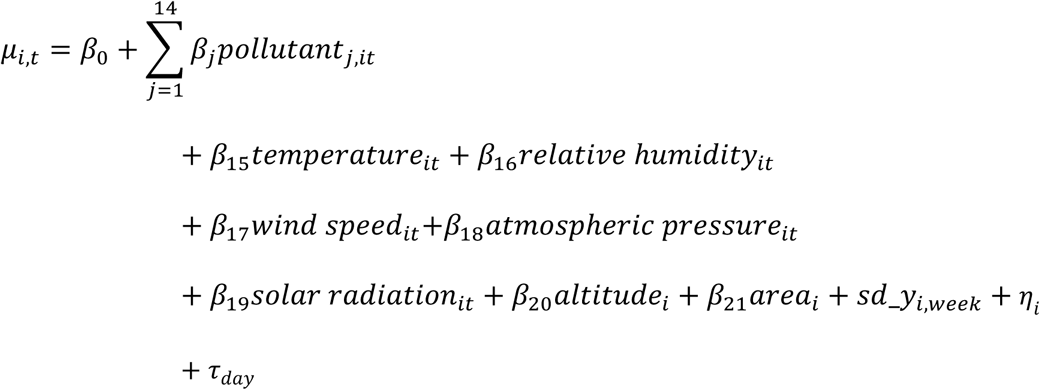

where i denoted the air pollution monitoring station where the pollutant was observed (i=1,2,…143); t was the time unit (month in the case of long-term exposure, day in the case of short-term exposure); *μ_i,t_* = *E*(*y_it_*), *y_it_* denoted the air pollutants of interest, PM_10_, NO_2_, O_3_ and PM_2.5_; *pollutants_j,it_* corresponded to the pollutant j measurements at station i and time unit t. Pollutants considered were, first, the pollutants of interest other than the pollutant for which the spatial prediction was made and, second, the rest of the pollutants (i.e., NO, SO_2_, CO, C_6_H_6_, H_2_S, Cl_2_, mercury, arsenic, nickel, cadmium and lead); *area_i_* was the area of the ABS i; *sd_y_i,._*, *η_i_* and *τ* denoted random effects.

In the models, we included *sd_y_i, year_*, *sd_y_i,week_* structured random effects, indexed on a standard deviation of the air pollutant that was being predicted, in the ABS i, during a particular year (2011 to 2018) and a particular week of 2020 (weeks 1 to 37), respectively. We chose, a random walk of order one (rw1) as the structure of the random effect. In the integrated nested Laplace approximations (INLA) approach (Rue *et al*., 2009, 2017), the random walk of order 1 for the Gaussian vector x is constructed assuming independent increments (R INLA project, 2021a):

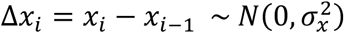

Following the INLA approach, when, as in our case, the random effects are indexed on a continuous variable, they can be used as smoothers to model non-linear dependency on covariates in the linear predictor. *η_i_* denoted a random effect indexed on the air pollution monitoring station. This random effect was unstructured (independent and identically distributed random effects) and captured individual heterogeneity, that is to say, unobserved confounders specific to the station and invariant in time.

We also included *τ_month_* and *τ_day_*, structured random effects indexed on time, in order to control the temporal dependency associated to possible seasonal effects throughout the year (long-term exposure) and throughout the week (short-term exposure). In this case, a model for seasonal variation with periodicity *m* (12 for long-term exposure, seven for short-term exposure), for the random vector (x_1_, x_2_,…,x_n_) (n>m) was obtained assuming that the sums were independent Gaussian with a precision Y. The density for x is derived from the n-m+1 increments (R INLA project, 2021b):

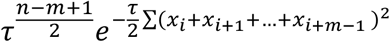

### 2.3. Inference

Inferences for GMRFs were made following a Bayesian perspective, using the INLA approach (Rue *et al*., 2009, 2017).

We started from the SPDE representation, which uses a finite element representation to define the Matérn field as a linear combination of basis functions defined on a triangulation of the domain (mesh, hereinafter). This consists of subdividing the domain into a set of non-intersecting triangles meeting in, at most, a common edge or corner (Lindgren *et al.,* 2011; Cameletti *et al.,* 2013).

Then, instead of projecting the subsequent mean of the random field onto mesh nodes to target locations where we do not have observed data, we performed the spatial prediction of the random field jointly with the parameter estimation process. For this, we projected the mesh into those locations with no air pollutants observed and then we jointly computed the posterior means at all the locations (with observed and unobserved air pollutants measurements) (Krainski *et al.,* 2020).

We separately estimated each year (long-term exposure) and each week (short-term exposure) and then merged every year and every week.

We used priors that penalize complexity (called PC priors). These priors are robust in the sense that they do not have an impact on the results, and furthermore, they have an epidemiological interpretation (Simpson *et al.,* 2017).

All analyses were carried out using the free software R (version 4.0.3), through the INLA package (Rue *et al*., 2009, 2017; R INLA project, 2021c). The maps were represented using the *leaflet* package (Cheng *et al.,* 2019).

### 2.4. Measures of predictive performance

The predictive performance of each model was assessed by cross-validation, considering a training set (2011 to 2018 for long-term exposure, weeks 1 to 36 - January 1 to September 8, 2020 -, for short-term exposure) and a test set (2019 for long-term exposure and weeks 37 - September 9 - to 48 - November 29, 2020- for short-term exposure).

The prediction accuracy was assessed by:

- Mean absolute percentage error (MAPE)

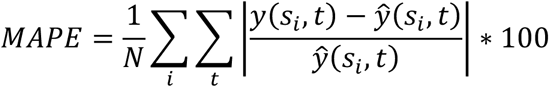

where N was the total number of available observations in the test set; *y*(*s_i_,t*) were the pollutant measurements (at station i and time unit t) at the test set; and 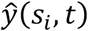 were the posterior means.

- Root mean square error (RMSE)

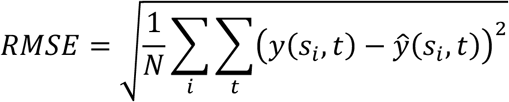

- Correlation coefficient

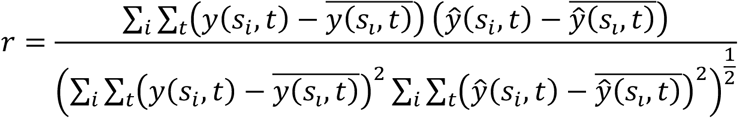

- Actual coverage of the 95% prediction intervals

### 2.5. Sensitivity analysis

We conducted two sensitivity analyses. In the first place, we carried out a new cross-validation and, in the second place, we changed the spatiotemporal model. In both cases, we consider the spatial prediction of long-term exposure to NO_2_.

As regards cross-validation, we considered, as training sets, five random samples from the monitoring stations in which NO_2_ was measured during the entire period 2011-2019. Specifically, we considered random samples of, approximately, 75% of the stations (58 out of a total of 77 stations), of 70% (55 stations), of 50% (41 stations), of 45% (35 stations), and of 20% (18 stations). As a test set, we considered the rest of the stations (19, 22, 36, 42 and 59 remaining stations, respectively).

Next, we calculated the measures’ prediction accuracy (explained previously).

With respect to the spatiotemporal model above, we considered an independent in time Gaussian field (GF), following Camelleti *et al*. (2013) we assumed a spatiotemporal Gaussian field that changes in time according to an autoregressive of order one (AR(1)).

Returning the measurement equation {2}:

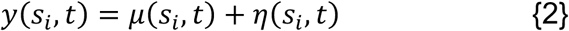

the realization of the spatiotemporal process, *η*(. , . ), was specified as,

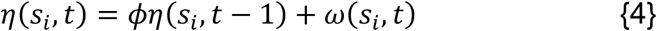

where |*ϕ*| < 1.

Here, it was *ω*(*s_i_,t*) what was assumed to be zero mean Gaussian and a Matérn covariance function:

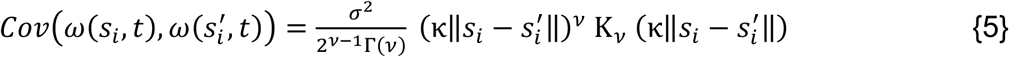

In the addition, in the GLMM specification of the large-scale component, *μ*(. , . ), in the linear predictor we included structured random effects indexed on year, *τ_year_*, in order to capture the long-term trend.

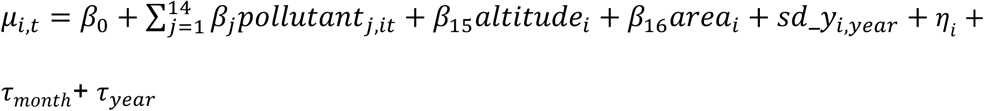

With this analysis, our objective was to compare not only the predictive performance of the model {1-2}, {4-5} with the one specified above {1-3}, but, above all, to compare the computation time in the inference of both specifications.

## 3. Results

Descriptive results are shown in Table 1. Regarding long-term exposure, we observed that, with the exception of O_3_, the daily averages of pollutants decreased in 2019 (PM_2.5_ 22.39% less, NO_2_ 12.88% less and PM_10_ 8.48% less).

In contrast, the daily average of O_3_ increased by 3.97% in 2019 compared to 2011-2018. With regard to short-term exposure, the levels of NO_2_ and of the PM_10_ were higher from September 9 (week 37) (23.11% and 4.87%, respectively). Conversely, the levels of O_3_ and of PM_2.5_ (although in this case only measured in three stations) were lower than the levels before September 9 (19.34% and 9.81%). When we excluded the lockdown (which took place in Spain from March 14 - week 11- to June 21 - week 25 -, both 2020), the variation from September 9 changed sign for PM_10_, it was 7.67% lower, they were moderated for NO_2_ (which was 3.64% higher) and O_3_ (12.48% lower), while they were increased in the case of PM_2.5_ (13.74% lower).

The measures of predictive performance are shown in Table 2. With the exception of PM_2.5,_ the results for long-term exposure were quite good. Achieved coverages of the 95% credibility intervals for predictions were greater than 90%, correlation coefficients were greater than 0.80, and MAPEs less than 10%. Furthermore, if Table 2 is compared with Table 1, it is observed that the reduction in the variability of the spatial prediction, measured between the ratio of the RMSE and the standard deviations of the pollutants observed, was, at most, one third of the standard deviations of the pollutants during the period 2011-2018 (19.40% for NO_2_, 25.71% for O_3_ and 33.89% for PM_10_), again with the exception of PM_2.5_ (the RMSE in this case was 59.92% of the standard deviation in the period 2011-2018). Therefore, except for PM_2.5_, our method managed to significantly reduce the variability of the spatial prediction around fairly accurate predictions. Although quite good, note that, in relative terms, the results for PM_10_ were somewhat worse than for gaseous pollutants (NO_2_ and O_3_).

**Table 2.** Measures of predictive performance. Spatiotemporal process independent in time Gaussian field.

**Long-term exposure**
|  | **MAPE** | **RMSE** | **Correlation** | **Coverage** |
| --- | --- | --- | --- | --- |
| **PM<sub>10</sub>** | 7.429% | 2.640 | 0.838 | 0.917 |
| **NO<sub>2</sub>** | 4.345% | 2.930 | 0.937 | 0.973 |
| **O<sub>3</sub>** | 6.795% | 5.287 | 0.916 | 0.945 |
| **PM<sub>2.5</sub>** | 16.037% | 3.068 | 0.696 | 0.750 |

**Short-term exposure – Training set Weeks 1-37**
|  | **MAPE** | **RMSE** | **Correlation** | **Coverage** |
| --- | --- | --- | --- | --- |
| **PM<sub>10</sub>** | 16.373% | 6.582 | 0.396 | 0.445 |
| **NO<sub>2</sub>** | 17.274% | 8.020 | 0.796 | 0.849 |
| **O<sub>3</sub>** | 20.604% | 9.711 | 0.726 | 0.837 |
| **PM<sub>2.5</sub>** | 49.897% | 6.366 | 0.398 | 0.081 |

**Short-term exposure – Training set Weeks 1-10 and 26-36 (excluding lockdown)**
|  | **MAPE** | **RMSE** | **Correlation** | **Coverage** |
| --- | --- | --- | --- | --- |
| **PM<sub>10</sub>** | 14.412% | 6.091 | 0.462 | 0.493 |
| **NO<sub>2</sub>** | 7.521% | 5.811 | 0.894 | 0.905 |
| **O<sub>3</sub>** | 9.233% | 7.192 | 0.853 | 0.890 |
| **PM<sub>2.5</sub>** | 39.936% | 5.589 | 0.398 | 0.081 |

MAPE: Mean absolute percentage error
RMSE: Root mean square error
Correlation: Correlation coefficient
Coverage: Actual coverage of the 95% prediction credibility intervals

The poor results obtained for PM_2.5_ are because of its smaller sample size. Although it is true that in the period as a whole up to 42 stations measured PM_2.5_, the year with the lowest number of active stations was 2018 (31 stations), with the rest of the years ranging between 33 and 35 active stations. No year fell below 40 active stations for the rest of pollutants (the year with the lowest number of stations measuring PM_10_ was 2018 with 94 stations, while the other years ranged between 100 and 107 stations; in the cases of NO_2_ and O_3_ it was 2015 with 59 and 44 stations, respectively, with the other years oscillating between 62 and 66, and 45 and 57, respectively).

Regarding the short-term exposure, first, predictive performance was worse when we did not exclude the lockdown period (which took place in Spain from March 14 to June 21, 2020) than when we did. In fact, note that predictive performance measures were much better for gaseous pollutants (NO_2_ and O_3_). The results for the coarse particles, PM_10,_ were quite poor (we did not interpret the results for PM_2.5_ as it was measured in only three stations). This was likely due to the lower number of stations where PM_10_ was measured (36 stations, versus 67 for NO_2_ and 50 for O_3_, see Table 1). The variability of the spatial prediction was reduced much less than in long-term exposure, especially for NO_2_. The RMSEs were between 33.83% for O_3_ and 43.49% for NO_2_, of the standard deviations of the pollutants (excluding lockdown).

The results of the sensitivity analyses, when the number of stations in the training set was greater than 40 and when the spatiotemporal Gaussian field changed in time according to an AR(1) (model {1-2}, {4-5}), were quite similar to the results for the spatiotemporal process independent in time Gaussian field (model {1-3}) and all the stations were included in the training set (Table 3).

**Table 3.** Sensitivity analyses. Measures of predictive performance.

**Spatiotemporal process independent in time Gaussian field**
|  | MAPE | RMSE | Correlation | Coverage |
| --- | --- | --- | --- | --- |
| Training set 58 monitoring stations (75%) | 4.973% | 3.205 | 0.930 | 0.963 |
| Training set 55 monitoring stations (70%) | 5.902% | 6.464 | 0.851 | 0.925 |
| Training set 41 monitoring stations (50%) | 6.072% | 7.201 | 0.786 | 0.870 |
| Training set 35 monitoring stations (45%) | 8.020% | 8.088 | 0.776 | 0.633 |
| Training set 18 monitoring stations (20%) | 8.806% | 10.159 | 0.609 | 0.590 |

MAPE: Mean absolute percentage error
RMSE: Root mean square error

**Long-term exposure**
|  | MAPE | RMSE | Correlation | Coverage |
| --- | --- | --- | --- | --- |
| NO<sub>2</sub> | 8.843% | 3.388 | 0.957 | 0.958 |

When we varied the number of stations in the training set, but used every year (2011 to 2019), the predictive performance seemed to depend on the size of the sample. The more stations the training set had, the better the results, while dramatically deteriorating with a small sample size. In particular, the cut-off appears to be 40 stations. Below this, the predictive performance measures were poor.

Although the predictive performance of the spatiotemporal Gaussian field model changed in time according to an AR (1) (model {1-2}, {4-5}) was very similar to that of the spatiotemporal process independent in time Gaussian field (model {1- 3}) (perhaps somewhat worse, in relative terms), the computation time was much longer. Using a 6-core Intel Core i9 (2.9 GHz 32 GB RAM), while the model inference {1-3} required on average 0.05 seconds per observation (a total of 569 seconds on average), the model {1-2}, {4-5} required 0.354 seconds (a total of 3,947 seconds), that is, seven times more computing time.

The maps of the posterior means and the posterior standard deviations for 2019 (in quintiles) of the spatiotemporal process independent in time Gaussian field (model {1-3}) for the long-term exposure of PM_10_, NO_2_ and O_3_ are shown in Figures 3. We decided not to represent the posterior means for PM_2.5_ because of its poor predictive performance. The spatial distributions of the subsequent means of PM_10_ and NO_2_ were quite similar, although in the high levels of NO_2_, (fourth and fifth quintiles) there was somewhat more spatial variation. Note that, unlike PM_10_ and NO_2_, the lowest levels of O_3_ (first and second quintiles) occurred in the urban areas. As expected, the uncertainty, as measured by the posterior standard deviations, was, in general, higher in those areas with few (or no) monitoring stations. Note, however, that higher levels of air pollutants do not always coincide with higher standard deviations.

**Figure 3a.**
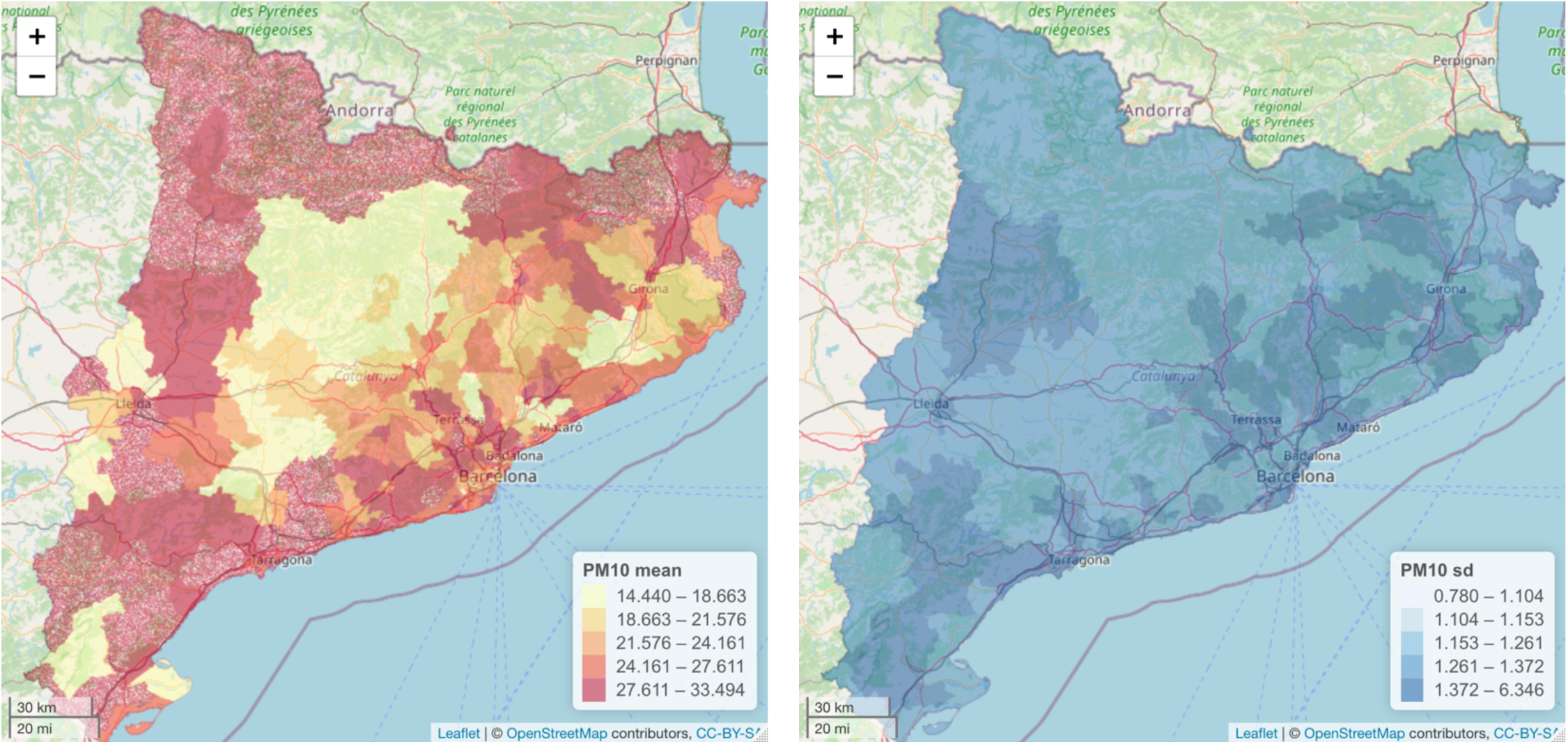
**Posterior mean and posterior standard deviation of PM_10_ for 2019. Spatiotemporal process independent in time Gaussian field**

**Figure 3b.**
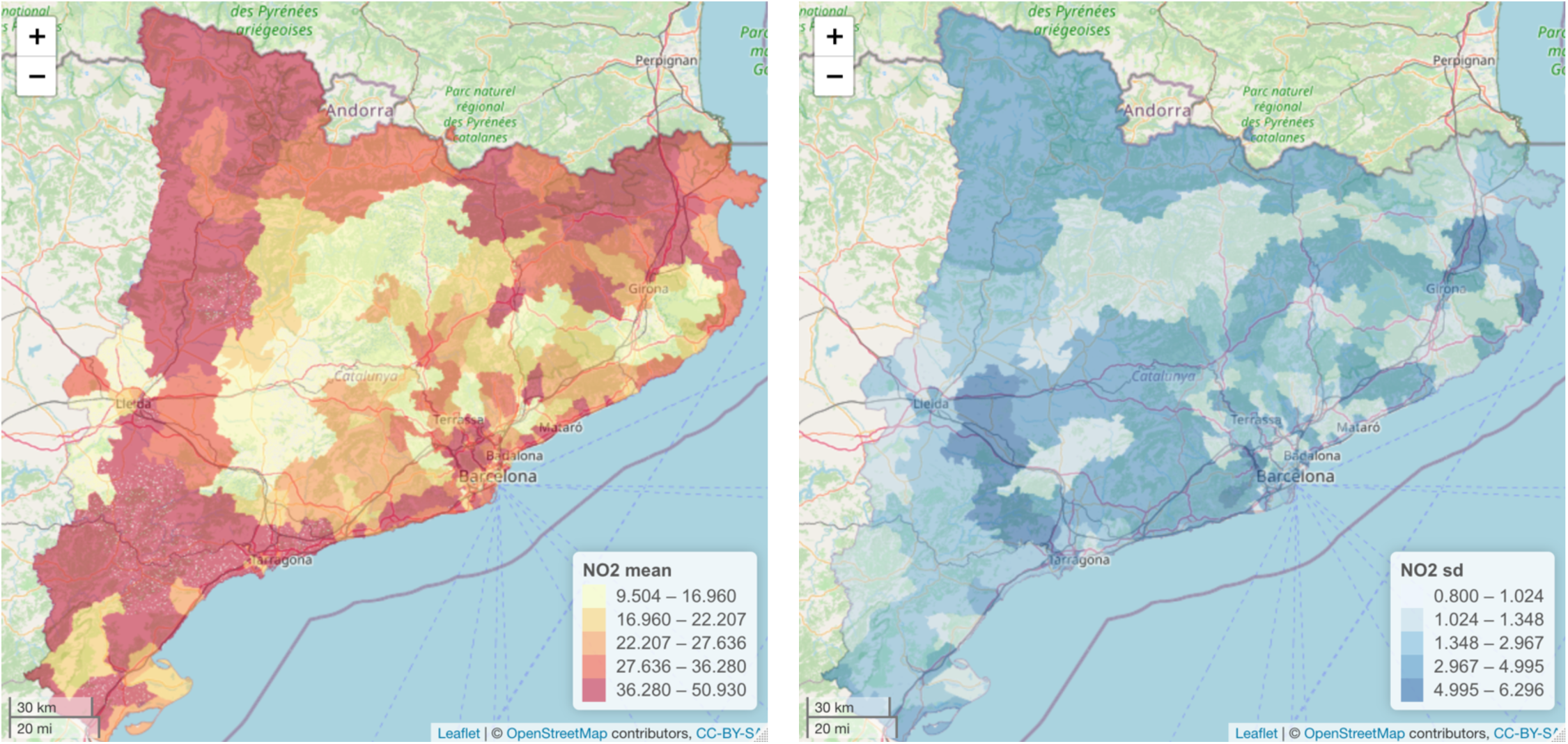
**Posterior mean and posterior standard deviation of NO_2_ for 2019. Spatiotemporal process independent in time Gaussian field**

**Figure 3c.**
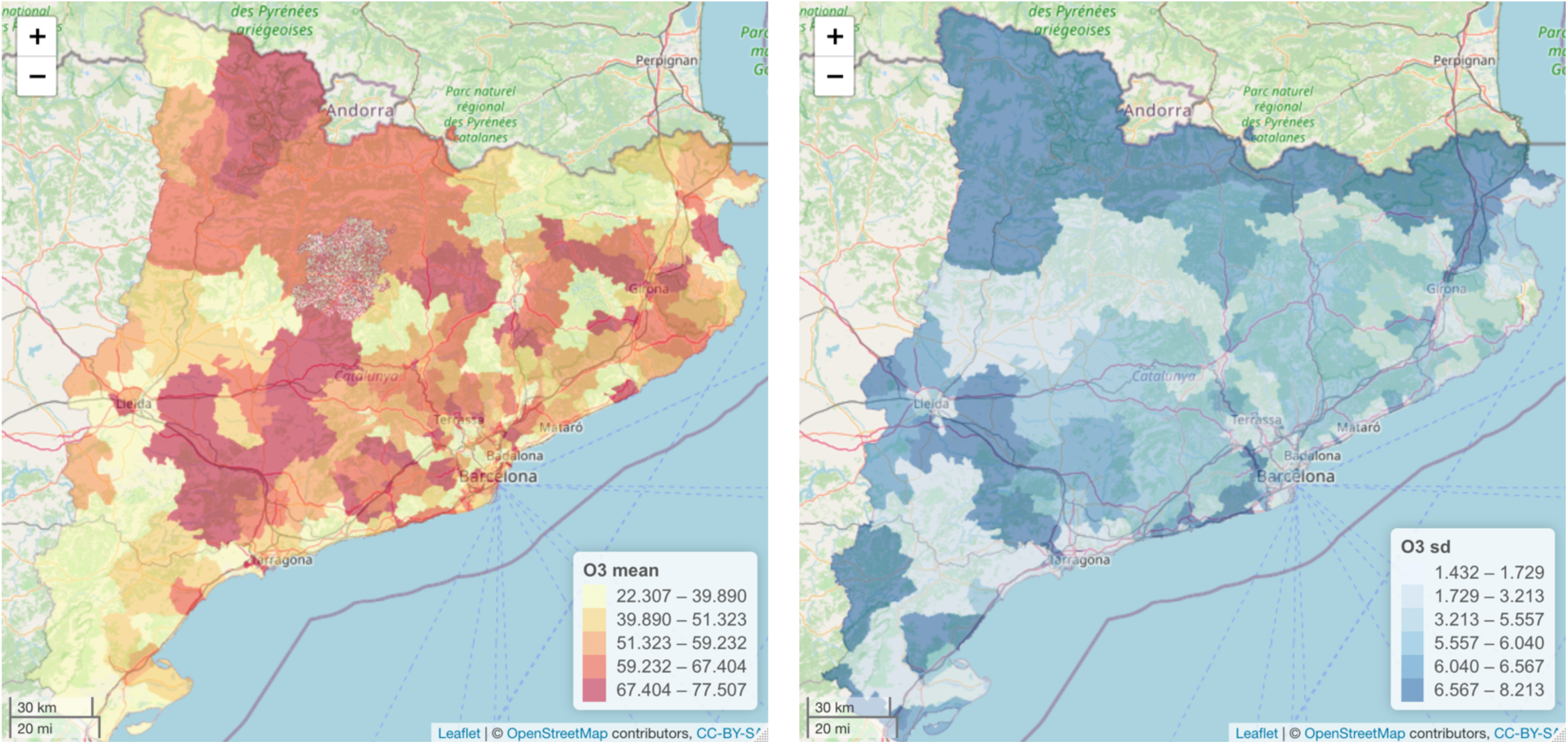
**Posterior mean and posterior standard deviation of O_3_ for 2019. Spatiotemporal process independent in time Gaussian field**

## 4. Discussion

Our results were quite good in terms of predictive performance, at least for those pollutants that were observed in more than 40 collecting stations (PM_10_, NO_2_ and O_3_ in long-term exposure and NO_2_ and O_3_ in short-term exposure).

The current coverage of the spatial predictions of these pollutants are in line with similar studies. Using the same model and the same data (PM_10_), but using two different methods for the inference, Camelleti *et al*. find coverage between 0.95 and 0.97 (using MCMC) (Camelleti *et al.,* 2011) and 0.897 (using INLA SPDE) (Camelleti *et al.,* 2013). Mukhopadhyay and Sahu (2018) find coverage between 0.91 and 0.92 for the spatial predictions for O_3_ (in our case, 0.89 for short-term exposure and 0.945 for long-term exposure), between 0.89 and 0.90 for PM_10_ (in our case, 0.917 for long-term exposure) and between 0.95 and 0.965 for NO_2_ (in our case, 0.905 for short-term exposure and 0.963 for long-term exposure). Note: we have preferred not to comment on the results in which we found poor predictive performance. Our coverage could also be comparable to those provided by Pirani *et al*. (2013) for the spatial predictions for PM_10_ (between 0.87 and 0.93), although it should be noted that these show the coverage at 90%.

The correlation coefficients between the observed levels of air pollutants and the subsequent means of the spatial predictions were higher in our case than in Camelleti *et al*. (0.863 when the inferences were made with MCMC - Camelleti *et al.,* 2011- and 0.702 when they were made with INLA SPDE - Camelleti *et al.,* 2013-, compared to 0.917 in our case), and than in Pirani *et al*. (2013) (between 0.73 and 0.78), in both cases for PM_10_. However, they were somewhat lower than in Mukhopadhyay and Sahu (2018) (0.88-0.89 for PM_10_, 0.92-0.94 for NO_2_, and 0.93-0.94 for O_3_). It should be said, nonetheless, that the number of observations in Mukhopadhyay and Sahu range between 56,625 (for PM_10_) and 100,138 (for NO_2_), while in our case we had 11,157 observations.

The reduction in the variability of the spatial prediction, can only be compared with Mukhopadhyay and Sahu (2018), since they are the only ones who show these standard deviations. In this sense, both Mukhopadhyay and Sahu and ourselves achieved a similar reduction in the variability of the spatial prediction.

Although good, the results of the predictive performance were less so for the spatial prediction of long-term exposure to PM_10_ (although it was being observed in the largest number of collecting stations, see Table 1) and for the short-term exposure for gaseous pollutants (NO_2_ and O_3_).

Regarding the spatial prediction of long-term exposure to PM_10_, we believe that it is a consequence of the location of the monitoring stations. The stations that measure PM_10_, although more abundant in urban areas, are also located in rural areas, while those that measure NO_2_ are located almost exclusively in urban areas. In the city of Barcelona, while 13% of NO_2_ is generated outside the municipality, it is 71% in the case of PM_10_ (Barcelona City Council, 2021; Saez *et al.,* 2020). It is not unreasonable to suppose that these figures can be extrapolated to the entire Barcelona Metropolitan Area, which comprises 41.75% of the total population of Catalonia and where the majority of PM_10_ and NO_2_ monitoring stations are located. In other words, while NO_2_ monitoring stations measured almost all NO_2_ pollution, PM_10_ monitoring stations did not collect all PM_10_ pollution data. This could also explain why the posterior means of the PM_10_ predictions exhibited less spatial variability than the NO_2_ predictions (Figures 3a and 3b).

With regard to the spatial predictions of short-term exposure, the reduction in the number of monitoring stations during 2020 could have led to a deterioration in the predictive performance. However, we believe it could also be due to the data behaviour during 2020. As a consequence of the lockdown to flatten the COVID- 19 pandemic curve, mobility was greatly reduced in 2020. Specifically, mobility was reduced by 40% on average, compared to pre-COVID-19 levels, during the lockdown and did not fully recover in the September-November 2020 period (being 5 to 15% lower, depending on the area of Catalonia [26]). We are sure that this anomalous behaviour would have influenced the predictive performance of the spatial predictions of short-term exposure.

The predictive performance of our model depends on the number of stations where pollutants are measured. We have found that with less than 40 stations, probably spread throughout the territory (although not necessarily homogeneously), the predictive performance deteriorates considerably.

Our method is quite similar to that of Camelleti et al. (2013). However, as we show with the sensitivity analysis, our method, in which we perform the inference year by year (or week by week) and then merge the subsequent ones, has a much shorter computation time, in addition to somewhat better results, even for such an atypical year as 2020.

Nevertheless, we are convinced that our results might not be as good if the spatial and temporal dimensions were dependent and not separable, that is, if the spatial dependence varied over time. Fortunately, the spatial dependence of air pollutants does not vary over time. Even during 2020, although air pollution levels decreased as a consequence of the reduction in mobility, the spatial dependence was more or less similar to previous years. For spatial dependence to vary over time, major changes in infrastructures or, likewise, lasting limitations in mobility that were not homogeneous throughout the territory, have to be produced. Of course, other types of spatiotemporal data could imply other results.

## 5. Conclusion

In this work, we have shown a hierarchical Bayesian spatiotemporal model that has allowed us to make fairly accurate spatial predictions with a low computational cost. Our model provides predictions of both long-term and short-term exposure. The only requirements of the method that we propose lie in a minimum number of stations being distributed throughout the territory where the prediction is to be made and that the spatial and temporal dimensions are either independent or separable.

## Data Availability

We used open data with free access using these sources.
Air pollutants
Departament de Territori i Sostenibilitat, Generalitat de Catalunya [Available at: https://analisi.transparenciacatalunya.cat/en/Medi-Ambient/Qualitat-de-l-aire-als-punts-de-mesurament-autom-t/tasf-thgu, last accessed on March 14, 2021].
Meteorological variables
METEOCAT, Generalitat de Catalunya. Meteorological data from XEMA [Available at: https://analisi.transparenciacatalunya.cat/en/Medi-Ambient/Dades-meteorol-giques-de-la-XEMA/nzvn-apee, last accessed on March 14, 2021].
AEMET. AEMET Open Data [in Spanish] [Available at: http://www.aemet.es/es/datos_abiertos/AEMET_OpenData, last accessed on March 14, 2021].
Digitized cartography of the ABS
Departament de Salut. Cartography [Available at: https://salutweb.gencat.cat/ca/el_departament/estadistiques_sanitaries/cartografia/, accessed on March 14, 2021].
Code will be available at www.researchprojects.es

http://www.researchprojects.es

## Acknowledgements

This study was carried out within the ‘Cohort-Real World Data’ subprogram of CIBER of Epidemiology and Public Health (CIBERESP).

## Ethics

Not applicable.

